# DYNAMICS OF THE COVID-19 PANDEMICS: GLOBAL PATTERN AND BETWEEN COUNTRIES VARIATIONS

**DOI:** 10.1101/2020.07.20.20155390

**Authors:** J. Krywyk, W. Oettgen, M. Messier, M. Mulot, A. Ugon, L. Toubiana

**Affiliations:** Well&Wiz, Paris, F-75001, France; GMAP SAS (Global Medical Affairs &Partners), Mulhouse, F-68100, France; Laboratory of Soil Biodiversity, University of Neuchâtel, 2000 Neuchâtel, Switzerland; ESIEE-Paris, Cité Descartes BP99, Noisy-le-Grand, 93162, France; Sorbonne Université, Université Paris 6, Sorbonne Paris Cité, F-75005, France; Sorbonne Université, Université Paris 13, Sorbonne Paris Cité, INSERM, UMR_S1142, LIMICS, F-75006, France; IRSAN, Paris, F-75008, France; INSERM, UMR_S1142, LIMICS, F-75006, France

## Abstract

The COVID-19 pandemic affected 203 countries between December 2019 and July 2020. The early epidemic “wave” affected countries which now report a few sporadic cases, achieving a stable late phase of the epidemic. Other countries are beginning their epidemic expansion phase. The objective of our study is to characterize the dynamics of the COVID-19 spread.

Data science methods were applied to pandemic, focusing on the daily fatality in 24 countries with more than 2,000 deaths, our analysis kin the end retaining 14 countries that have completed a full cycle.

The analysis demonstrates a COVID-19 dynamic similar in these studied countries. This 3-phase dynamic is like that of common viral respiratory infections. This pattern, however, shows variability and therefore specificity which the method categorizes into clusters of “differentiated epidemic patterns”. Among the 5 detected clusters, 2 main ones regroup 11 of these countries, representing 65% of the world deaths (as of June 24, 2020).

The pattern seems common to a very large number of countries, and congruent with that of epidemics of other respiratory syndromes, opens the hypothesis that the COVID-19 pandemic would have developed its “natural history” by spreading spontaneously despite the measures taken to contain it. The diversity highlighted by the classification into “formal clusters” suggests explanations involving the notion of demographic and geographic epicenters.

## Introduction

The first SARS-COV-2 patients were detected around a food market in the city of Wuhan, in China, in December 2019 [1]. The outbreak then spread rapidly to all countries. It reached Europe at the end of February 2020. To date, the COVID-19 pandemic has struck most countries in the world: 203 countries have registered at least 1 case. The pandemic continues to expand geographically, with very high impacts in the Americas but also in some countries, such as India, which had been spared so far.

Each country starting on different dates illustrates various epidemic phases. Countries that were affected at the beginning of the pandemic (China and many European countries) consider a completed epidemic cycle, while in other countries are in their early phases.

The availability of data on the evolution of the pandemic is one of the remarkable features of this global event. This data was available quickly and has been compiled in unique sources, updated daily with a fairly good accuracy and relative completeness for each country [2]. While acknowledging some heterogeneous reporting policies, the daily data sets present the advantage of reflecting real world dynamics of the epidemic, over time and in different places.

Modern data science techniques can characterize and compare temporal data. It seemed relevant to apply these methods to the COVID-19 pandemic. Among the available indicators, the evolution of deaths was considered most reliable to compare across countries.

A preliminary analysis of the evolution of mortality selecting only countries with a complete epidemic cycle - determined a general pattern of COVID-19 similar to some common winter respiratory viral infections. Thus, COVID-19 appears to reproduce a three-phase pattern: a phase of rapid progression of incidence (1) until a peak is reached (2) from which a decrease (3) begins, until a low point of stabilization where the cycle is considered to be over.

Although this pattern is rather universally verified, there is some variability in the dynamic behavior of COVID-19 between countries.

The objective of this analysis is to characterize the dynamics of the propagation of COVID-19. To do so, requires a rapid and robust methodology: countries were categorized according to “differentiated epidemic patterns” based on their similarities or dissimilarities in terms of epidemic dynamics. We hypothesized a dynamic variability in time and space. The goal was to test the hypothesis looking for a specific epidemic pattern for COVID-19, exploring the natural history of this pandemic independently of the measures taken to contain it.

## Methods

### Data

The data used are those available on the John Hopkins University website [2]. This university makes several indicators available to the general public in open access, including the daily cumulative number of new cases, deaths and cured individuals. However, although the WHO published a clinical definition of cases very early on, it has evolved over time in part due to the evolving diagnostic tests. There are caveats about the reliability of case counting and definition, the number of tests performed, and whether or not some cases (eg. from retirement homes) are even reported or defined in different countries. These limitations pose serious difficulties to compare the dynamics in terms of the number of infected people. This determined the choice to more robust indicators such as the “daily death” used in all our analyses.

### Analysis pipeline

The data have been subject to the following selection and specific processing operations:

• Selection
  ○ Only countries infected with more than 2000 COVID-19 deaths, representing the ninth decile (D9) were considered, i.e. a total of 24 countries.
  ○ Only countries that are advanced in the development of the epidemic, i.e. have passed the peak (stage 2) and are well into the phase 3 decline **[Figure 1]** have been retained.

**Figure 1:**
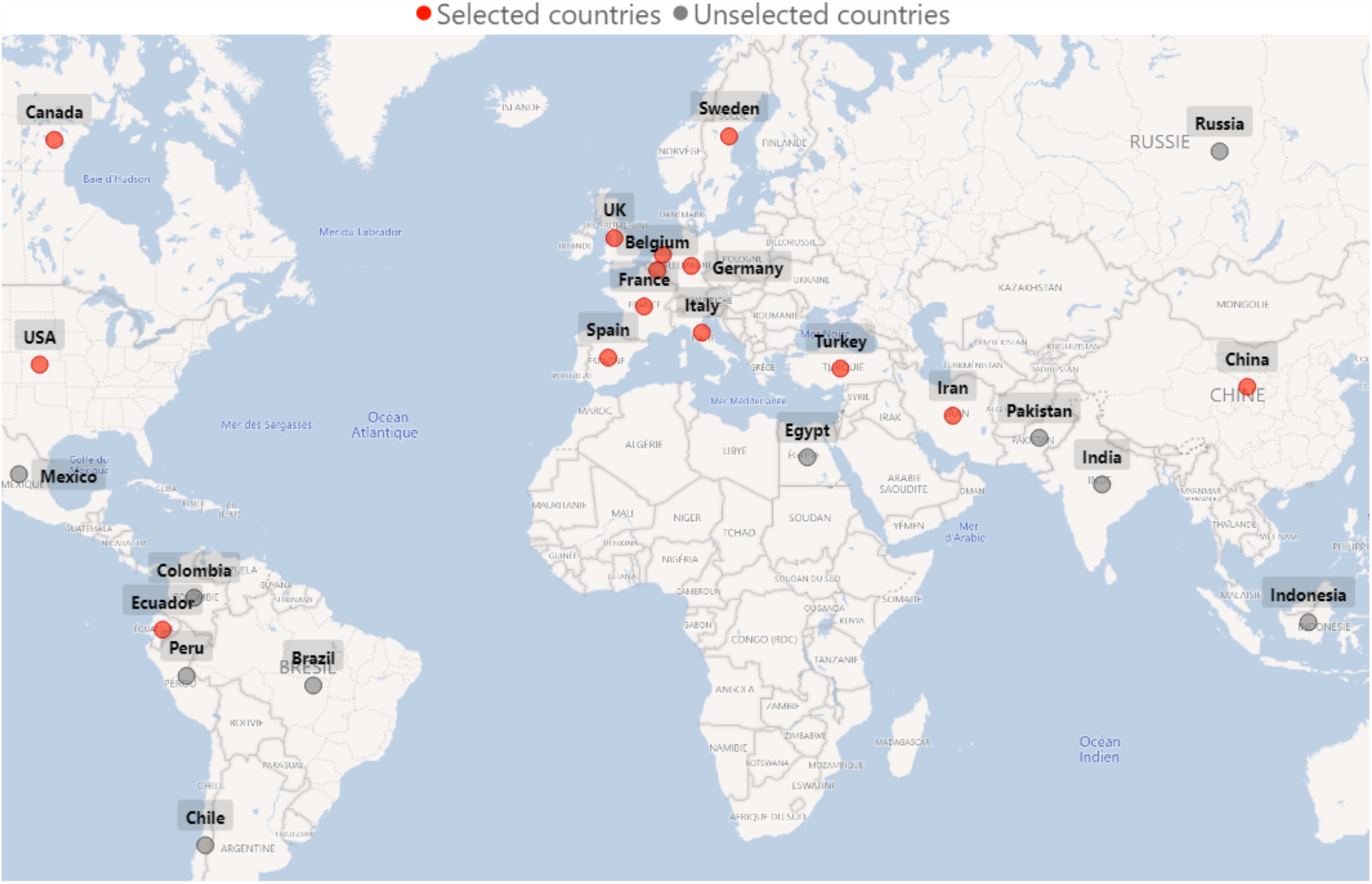
24 countries with a death count > 2000 as of 24 June 2020. The countries most affected by the COVID-19 epidemic can be found on all continents. The most advanced in the epidemic cycle, shown in red in the graph, are those that were affected earliest and chronologically, moving from East to West: China then Europe then North America. Analyses of the epidemic pattern are carried out on the subset of 14 countries for which the three phases of growth, peak and decline have been observed.
• Pre-processing
  ○ The curves for each of the selected countries were initially constructed using an n=1 day incidence
  ○ Deferred readjustment values (i.e. by catching up) are anomalies and replaced, where appropriate, by the average value of the two closest neighbors (n+1; n+2).
  ○ Negative impacts, corresponding to corrections for over-reporting of deaths, have been replaced by the average value of the two closest neighbors (n+1; n+2).
  ○ A double smoothing of the curves, in increments of 1, was performed using a triangular moving average (TMA). This smoothing, often used in other fields [3], removes background noise, smooth curves further and highlights trend. The TMA used here has two components: a 10-day moving average (SMA) and a 5-day moving average, which buffers the possible effects of under- or over-reporting observed on week-end data. SMA of size n for each Pi was computed as follow

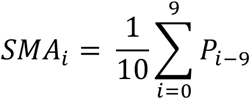

Where Pi is the value at date **i** and **n** the window size, in days. It follows that TMA of each point P is computed as:

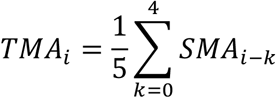
  ○ Time series were reduced centered [4] for each country to cancel the influence of a size effect during clustering.
  ○ All country curves were rescaled from a “start of the epidemic” as reaching a slope of 0.35 [5].
• Global pattern
  ○ Once pre-processed, the average of the curves for the selected countries was calculated, with a 95% confidence interval [Figure 3].

The study of time series is a very active field of research, this data format frequent and exploited, especially in acoustics [6], power management [7] or health [8]. In addition to prediction, time series can also be considered from a classification point of view, i.e. the grouping of curves according to their similarities/dissimilarities. The unsupervised classification approach makes it possible to apprehend logics and structures that differentiate behaviors [9]. For this purpose, and in many applications [10], the k-means algorithm [11] is used, together with the Euclidean distance function. Although the Euclidean distance is intuitive and simple, it is not optimal for time series of variable ranges.

Dynamic Time Warping Distance (DTW) [12] is suitable for comparing time series of different sizes. This distance, used in the k-means algorithm, gives more accurate results than the Euclidean distance [13]. The following section provides a general explanation of k-means clustering and DTW distance measurement.

### k-mean clustering

k-means clustering [11] is a well-known and very simple clustering algorithm. The principle consists in regroup similar data in the same cluster using an objective function that minimizes the sum of squared errors between a cluster center and its members.

The algorithm works as follows:

1. Initialization of the k cluster centroids
2. Measuring the membership between each data and all cluster centers and assigning the data to the appropriate cluster
3. Calculating a new cluster centroid for each cluster using an averaging function
4. Repeat steps 2 and 3 until the data in the clusters is stable

### Dynamic time warping

The DTW distance [12] is a measure of similarity that is generally used for time series, especially in classification [13]. The optimal alignment and distance measurement between two P and Q sequences can be determined as follows:

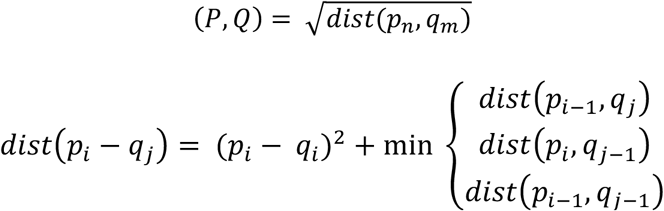

The DTW distance is used to measure the similarity between time series and calculate the relative cluster centers to optimize our clustering.

#### Optimal number of clusters

The optimal number of clusters was determined with the silhouette coefficient, a measure of the quality of the data segmentation. This was assessed by varying the number of clusters from 3 to 7, with a gamma parameter from 0.3 to 0.9, and by selecting the solution maximizing the silhouette score. The chosen solution, with a silhouette score of 0.37, segmented the countries into 5 clusters.

## Results

An examination of the dynamics of the epidemic revealed similarities and contrasts. Of the 24 countries considered (in the ninth decile), 14 are in an advanced stage of the epidemic cycle, having reached stage 3 of rapid decline (see above), the others are still in phase 1 of rapid growth and have therefore not yet passed the epidemic peak. The results selected, report on the subset of 14 countries which are highlighted in red **[Figure 2]**.

**Figure 2:**
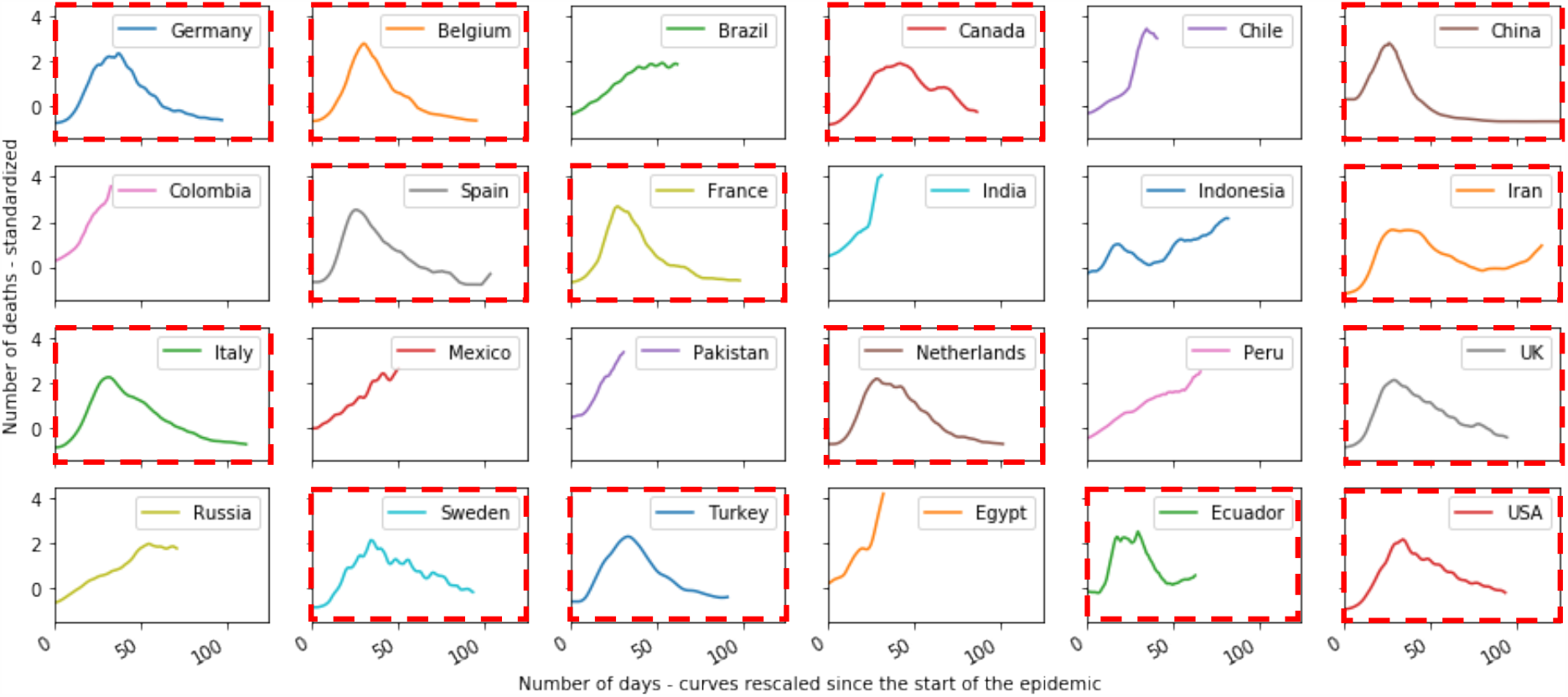
Epidemic dynamics for countries with a death count > 2000. Out of the 24 countries with significant numbers of deaths, only 14 of them - shown in red - have gone through a full cycle integrating the three characteristic phases of the epidemic. The other countries, insufficiently advanced in the cycle, are not taken into account for the purpose of comparing the shape of epidemic cycles.

**Figure 3:**
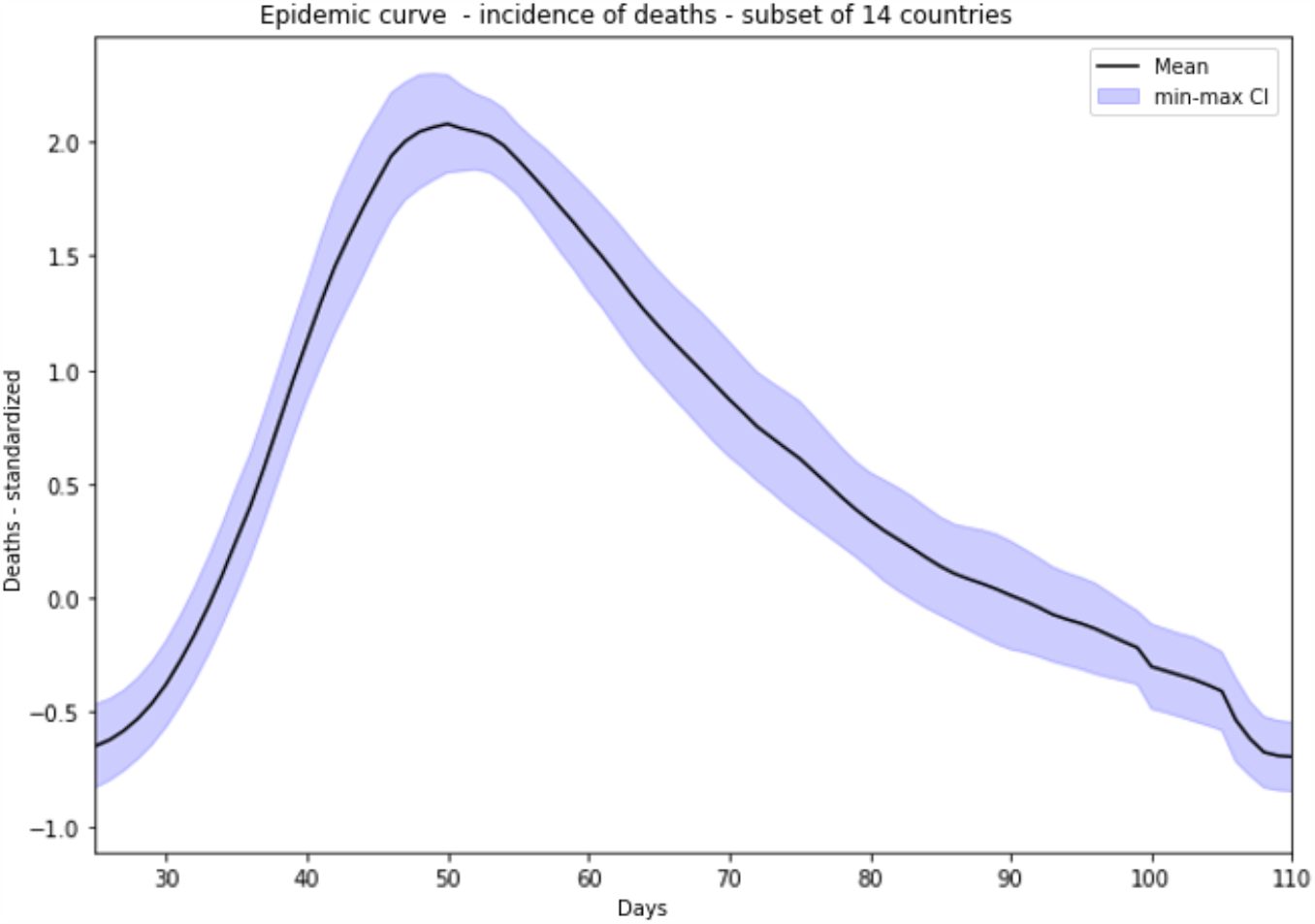
Averaged epidemic dynamics - incidence of deaths - for the 14 countries studied. The above curve represents the average profile of the 14 country curves, shifted to phase 1 growth from the reduced centred death data. A characteristic pattern consisting of 3 phases emerges: an exponential growth phase, a peak with a slightly extended plateau and a decrease phase with a relatively regular slope, more linear than the first phase.

The apparent similarity of the epidemic dynamics of the 14 selected countries are prone to construction of the average curve of the evolution of the number of deaths, covering a period of 110 days **[Figure 3]**. The global pattern can be clearly identified, consisting of 3 characteristic phases: strong growth over a period of 3 to 4 weeks, until a “peak” is reached, where the incidence of deaths gradually decreases, at a slower rate than the first phase of growth. This general pattern is therefore observed overall for all the countries under consideration.

A comparison of the 14 countries can be assessed using the epidemic curve classification **[Figure 4] [Figure 5]**.

**Figure 4:**
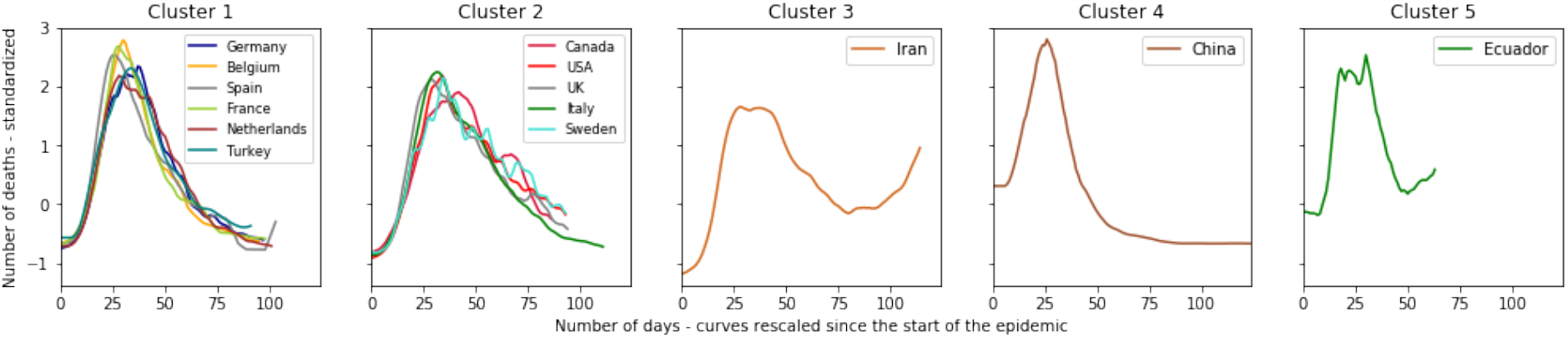
Clustering of the 14 countries into 5 classes based on the death curves, calculated as centred-scaled moving averages for each country. Behind the common general pattern, there are notable differences with 5 shapes of epidemic dynamics. Most European countries are grouped in cluster 1, with a profile very close to the average pattern. Cluster 2 brings together 5 more atypical countries, characterized by a phase of decreasing trend with an uneven rate. Iran, China and Ecuador each present singular epidemic dynamics.

**Figure 5:**
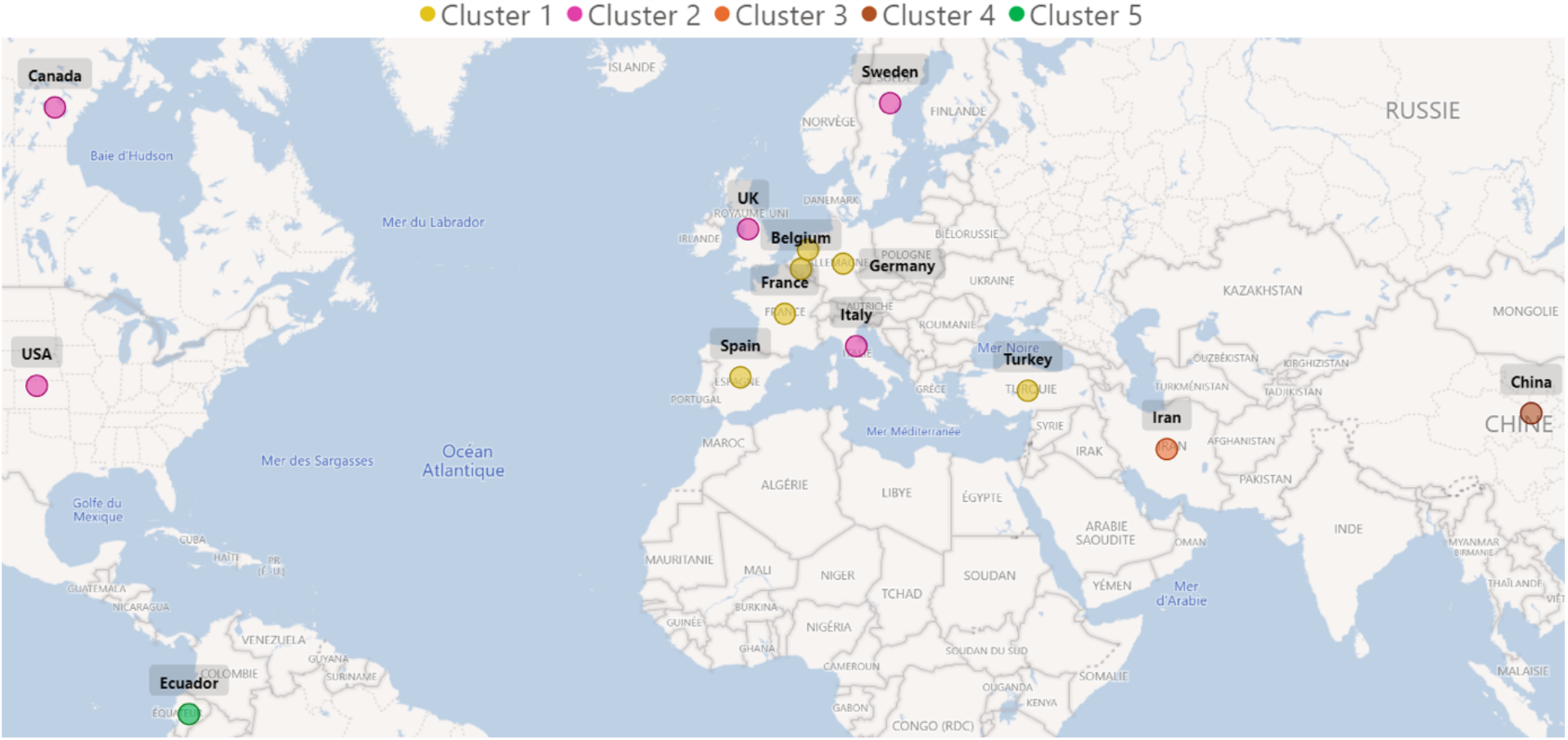
14 countries identified according to the clusters they belong to. The results of clustering highlight the geographical proximity of countries belonging to cluster 1. The two large territories (Canada, United States) are also grouped together within the same cluster, which also includes the United Kingdom, Sweden and Italy. The more isolated countries, far from the others, make up the remaining clusters.

We distinguish 5 clusters, with specific profiles:

1. Cluster 1 gathers 5 European countries (Germany, Belgium, Spain, France, the Netherlands) plus Turkey, with a very strong similarity of their epidemic dynamics: a high peak and a regular decreasing phase. Nevertheless, within this cluster, highlights Germany, the Netherlands and Turkey, with a slightly more crushed peak and therefore a slightly longer maintenance at this stage of transition.
2. Cluster 2 brings together 5 countries (Canada, United States, United Kingdom, Italy, Sweden) which share a common pattern: the decline phase is more uneven.
3. Cluster 3, exclusively made up of Iran, has a characteristic profile with a rebound from the 75th day.
4. Cluster 4, made up of China, has a very symmetrical curve profile between growth and decline phases, and a short time to plateau. Note the return to a (counter-intuitive) negative value due to the centered-reduced moving average,
5. Cluster 5 highlights the atypical nature of Ecuador: a plateau more spread out at the peak and a less advanced stage in the epidemic cycle.

## Discussion

The COVID-19 epidemic, like other viral diseases, has an unsurprising dynamic characterized by the 3 phases of growth, peak and decline. The incidence of deaths can be observed with a certain similarity between the countries that are most advanced in the epidemic, revealing a dynamic specific to COVID-19. Behind this common global pattern, however, lie differences from one country to another, which this work highlights by classifying into time series.

At this stage of the analysis, we can hypothesize which factors contribute to the differences between countries reflected in the classification. The notion of epicenter may play a role in the process of epidemic propagation, and consequently on the shape of the epidemic pattern, particularly in phase-3 of decline. An epicenter constitutes a locally active propagation hotspot.

- It should be noted that countries of relatively small surface area (Germany, Belgium, Spain, France, the Netherlands, Turkey), which have had a small number of epicenters, present an epidemic pattern characterized by a regular decrease, i.e. without marked rebounds. No doubt the epidemic control policies (lockdown, screening policies, etc.) may also have had an impact in limiting the multiplication of epicenters. In this respect, China reflects the case of a single controlled epicenter (city of Wuhan) and perhaps constitutes the basic pattern of COVID-19.
- Conversely, countries with a larger surface area (Canada, United States, Iran), involving a larger number of potential epicenters, are characterized by a dynamic with a more uneven phase of decline, i.e. with rebound phenomena probably linked to the appearance of successive epicenters. The cases of Russia and Brazil will be assessed closely and could - a hypothesis yet to be confirmed - ultimately present an epidemic pattern resembling that of the United States or Canada.
- Finally, the situation in the United Kingdom or Sweden, with a longer plateau phase, followed by a slower and relatively uneven decline, may relate to the specific containment policies in these countries.

This analysis will be updated when more country epidemic cycles are complete, which is not the case for many of the 203 affected countries. A study of epidemic patterns performed by epicenter could also refine previous hypotheses.

When such data become available, it will be useful to work on excess mortality attributable *a posteriori s*to COVID-19.

Finally, the variations observed through our analyses should be confronted with other explanatory hypotheses: can the different strains of the virus be associated with the general epidemic pattern? To what extent do the epidemic management policies also contribute to this pattern? How do the socio-geographical characteristics of the territory (surface area, urbanization poles, number and distance between epicenters, communication axes) affect the epidemic dynamics?

## Conclusion

The study addresses the epidemic form specific to COVID-19 and, its characteristics. To answer this question, techniques from the field of time series data science were applied. The upstream preparation of the data, drawing the shape of curves, and the subsequent classification of these dynamics, according to their similarities/dissimilarities, illustrates an epidemic form.

The dynamics of the COVID-19 epidemic must be considered as having a general pattern. Firstly, the average envelope of death curves, constructed from the 14 most affected countries (i.e. 70% of global deaths), shows a characteristic pattern. Secondly, within this envelope itself, there are dissimilarities that encourage segmenting the curves. The method used highlights two main clusters for 11 of these countries, representing 65% of the world’s deaths (as of 24/6/2020). The search for a history common to the countries of the same cluster resulting from the classification suggests geographical and demographic explanations involving the notion of epicenters contributing according to their numbers and their relationships within countries.

Ultimately, we wish to quantify the intrinsic characteristics of clusters, and to know how to describe and differentiate them objectively. In particular, we could explain the rebounds, visible in certain curves and invisible in the one we call the basic pattern of COVID-19, rebounds that we imagine result from the contributions of new epicenters, rather than from the resurgence of the Covid-19 epidemic itself.

## Data Availability

Data Available here : https://coronavirus.jhu.edu/map.html

https://coronavirus.jhu.edu/map.html

## Acknowledgements

This work was carried out within the Biotechnology Department of Well&Wiz, with the academic support of the Laboratoire d’informatique de Paris 6 and the Biotechnology Department of ESIEE-Paris.

